# Exploring the potential of geospatial mapping of emergency call data to improve ambulance services for older adults: a feasibility study in the South Central region of England

**DOI:** 10.1101/2025.03.06.25323481

**Authors:** Carole Fogg, Phil King, Vivienne Parsons, Nicola Dunbar, Marcel Woutersen, Julia Branson, Helen Pocock, Patryk Jadzinski, Chloe Lofthouse-Jones, Bronagh Walsh, Dianna Smith

## Abstract

**Background:** Ambulance Trusts across the UK serve vast and varied regions, impacting equitable healthcare access, especially for older patients facing urgent, non-life-threatening conditions. Detailed variation in demand and service provision across these regions remains unexplored but is crucial for shaping effective care policies and organisation. Geospatial mapping techniques have the potential to improve understanding of variation across a region, with benefits for service design.

**Aim:** To explore the feasibility of using geospatial techniques to map emergency 999 calls and outcomes for older adults within an academic-healthcare collaboration framework.

**Methods:** The study utilised administrative and clinical data for patients aged ≥65 who made urgent calls to a regional ambulance service within one year. This data, aggregated by geographical area, was analysed using geospatial software (ArcGIS) to create detailed chloropleth maps. These maps displayed metrics including population demographics, number of calls, response times, falls, dementia cases and hospital conveyance rates at the middle-layer super output area level. Feedback was solicited from internal stakeholders to enhance utility and focus on service improvements.

**Results:** The analysis unveiled significant regional disparities in emergency call frequencies and ambulance requirements for older adults, with notable variations in hospital conveyance rates, ranging from 22% to 100% across different areas. The geographical distribution of falls and dementia calls corresponded with the older population’s distribution. Response times varied by location. Stakeholders recommended additional data incorporation for better map utility and identified areas for service enhancement, particularly in addressing conveyance rate disparities for falls.

**Conclusions:** Leveraging aggregated ambulance service data for geospatial mapping of older adults’ care demand and provision proves to be both feasible and insightful. The significant geographical variances in hospital conveyance highlight the need for further research. The development of academic-healthcare partnerships promotes resource and sharing of expertise, which should substantially benefit patient care for this vulnerable group.

## Introduction

UK National Health Service (NHS) ambulance services are under intense pressure to deliver timely and safe care. Immediately life-threatening conditions have to be prioritised, and older people (aged ≥65 years) with falls and symptoms related to long-term chronic conditions with lower category calls may experience long delays before an ambulance arrives. Such patients often have recurring needs for emergency care due to lack of alternative care pathways or long waits for social care assessments. These patients may also have dementia, putting them at greater risk of clinical deterioration whilst waiting for their needs to be met, resulting in hospital admissions which further increase the risk of adverse outcomes, regardless of the reason for admission [1, 2]. Older adults with urgent or less urgent calls, which often include those living with dementia, are a key strategic focus for the development of alternative care pathways within the context of maturing integrated care systems and the drive to reduce conveyance to hospital and avoidable hospital admissions [3].

The NHS is a large organisation providing acute and preventive medical care that is free at the point of delivery in England and the devolved nations of the UK. When the NHS was set up in 1948, the focus was on treating single conditions or illnesses. However, population health and care needs have changed over time, with more people living longer and with comorbidities that require ongoing care. NHS services are delivered by multiple separate provider organisations, including primary healthcare practices, acute hospitals and ambulance services. As care has become more complex, the 2022 Health and Care Act aimed to make it easier for organisations to work together and join up services for patients, providing integrated care [4]. The NHS remains separate from the social care system, access to which is means-tested.

Ambulance services are a key part of NHS integrated care systems and cover wide geographical regions with large populations with a range of urbanisation and socio-demographic characteristics. Health and social care services within regions are variable in content and are not evenly distributed, so staff working in different locations within the region have to make decisions for onward patient care based on availability of localised services. These geographic patterns in population and care may impact on service demand and patient and service outcomes and further adds to the challenge of managing a patient group with already complex needs. Previous work has explored disparities in access to NHS services using geospatial methods or has assessed the feasibility of using geospatial data to identify priority areas for interventions [5, 6]. Such data are used to support decision making in local government and associated organisations such as Integrated Care Boards, and the visual nature of these data allow for ease of interpretation for non-experts. There are a few examples of research using geographic data using tracking devices for people with dementia and mapping prevalence rates, but examples of use to inform large-scale care and policy planning decisions are scarce [7]. However, geospatial projects in ambulance services have had significant impact, for include improving cardiac arrest response times and identifying vulnerable communities during Covid-19, and such techniques could also be applied to other patient groups such as those living with dementia [8, 9].

The South Central Ambulance Service (SCAS) region in England provides services to more than 4 million people and covers the counties of Berkshire, Buckinghamshire, Hampshire and Oxfordshire. Adults aged ≥65 represent a large proportion of the demand for services, comprising 17% of 111 calls (the NHS non-emergency phone service for urgent medical help or advice) and 48% of 999 calls (the emergency response service) - an average of 21,200 per month - whilst comprising 18% of the overall population [10]. An estimated 14% of emergency attendances are to adults living with dementia [11]. Following previous work to develop the scope for paramedics to identify and record dementia on the ambulance electronic patient record (ePR) [12, 13], a recent quality improvement project within SCAS has systematised recording of dementia [14, 15]. Improved recording of dementia in ambulance data provides an opportunity to explore factors associated with service demand associated with dementia, and to inform configuration of emergency and associated services to improve patient experience and outcomes, identify and address inequalities, and enable best use of existing resources.

We report on a project exploring the feasibility of using geospatial data to map lower category calls to adults aged ≥65, including a subgroup of those with dementia. This project was a collaboration between the ambulance trust and local academic partners within their operational region. The aim of the project was to establish whether, and how, the data can be extracted, mapped and presented in a way which is useful to the ambulance service and the wider regional health and care system, and to identify preliminary patterns in service use and outcomes whilst protecting patient confidentiality.

## Methods

### Objectives

The specific objectives of this project were to: (1) test feasibility and methods of data specification, extraction and aggregation from the SCAS data warehouse; (2) produce maps representing the extracted data to describe the population and service demand and outcomes; (3) discuss the data with clinical stakeholders to identify topics of interest and explore topics for further quality improvement or research.

### Formation of collaborative team

To manage the stages of the project, a collaborative team was formed which included clinicians, emergency care research experts, business intelligence leads and analysts and community first responder leads, together with academic geospatial analysts and health service researchers.

### Data specification, extraction and sharing

The team worked to understand and define data items for the study. This included administrative, socio-demographic, and clinical and incident datasets. Sources included records from the SCAS Computer Aided Dispatch system (CAD), the patient electronic patient record (ePR) and locations of hospitals, community first responder schemes and ambulance stations, in addition to publicly available census data and deprivation indices.

The core data were records (routinely collected and retrospective), of incidents for patients aged ≥65 seen by SCAS between April 2022 and March 2023 inclusive with incidents categorised as urgent (category 3 and 4). These were provided as an aggregated dataset, with the number and type of incidents aggregated by Lower Super Output Area (LSOA) level; no individual-level data were used. A data sharing agreement between organisations was signed, and a secure file sharing system used to transfer data.

### Data items used in the analysis

Four types of datasets were used in analyses:

i. Incidents and attendances for patients aged 65+ and 75+, average response times (interval between time of call and first clinical response), time on scene <1 hour or ≥1 hour, number with falls (also split by 65+ and 75+), top five presenting complaints, number conveyed to hospital, number of falls with conveyances.
ii. As (i) (from the attendance point on the pathway) for a subgroup of people with a record of dementia.
iii. 2011 LSOAs and Middle Super-Output Areas (MSOAs) (ONS Administrative Boundaries, 2011) in the region served by SCAS (Berkshire, Buckinghamshire, Hampshire, and Oxfordshire) with contextual data of population numbers from the 2021 Census [16], Indices of Multiple Deprivation (IMD) quintiles and Income Deprivation Affecting Older People’s Index (IDAOPI) quintiles [17].
iv. Locations of hospitals (Ordnance Survey Open Map) [18], ambulance stations and locations of community first responder schemes (SCAS).

### Data analysis and visualisation

ArcGIS software (www.arcgis.com/) was used to produce choropleth maps at LSOA (average population of 1641) and MSOA (average population of 7787) [19] level of the data items listed above. LSOAs and MSOAs covering the SCAS region were extracted and two layers were created from this. Contextual, incident and attendance data for patients (including dementia subset records) were joined using the common LSOA/MSOA code to be able to display this data according to LSOA/MSOA. Postcodes of the community first responders and ambulance stations were transformed into grid references. Hospital locations were extracted from OpenMap as points and polygons. Where locations were represented in polygon format, points were created and joined to the point dataset.

### Data interpretation

Two stakeholder sessions were held with representatives from SCAS, including staff working in different areas within the region, and those with responsibilities for dementia education and care, community engagement operations and patient pathways, in addition to clinical academic partners working with paramedics in the region (University of Portsmouth and Oxford Brookes University). The sessions focussed on: identifying key messages from the maps; considering how the maps could be amended or improved; how the maps might be used in planning services; which additional geospatial analysis could be undertaken; and the key messages/areas to explore with external stakeholders. Notes were taken throughout the sessions and a summary was produced which was circulated to attendees for amendments and approval.

### Public and patient involvement

This feasibility study is one of a group of projects looking at older people’s interactions with ambulance services. The study was informed by discussions with relatives and carers of older people with recent interaction with ambulance services, and by public volunteers with the ambulance service (Emergency Responders). The results will be discussed in a dissemination session with a wider group of public members in order to design further research.

### Ethics and governance

This project used routinely collected anonymised aggregated data therefore did not require ethical approval from the NHS Health Research Authority. The project was approved by the SCAS Clinical Review Group and the data sharing agreement signed in May 2023.

## Results

### Overview of data

The data represents 221,752 incidents (combined 111 and 999 calls) for people aged ≥65 were aggregated by MSOAs to enable more effective visualisation of the data. Of the incidents, 32% (n=71,545) were attended by a clinical team; of which 20% (n=14,636) were to people aged 65-74, and 80% (n=56,909) to people aged ≥75. Of the attendances, 52% (n=37,385) resulted in a conveyance to hospital or alternative service, of which 21% (n=7,881) were for people aged 65-74, and 79% (n=29,504) for people aged ≥75. Falls accounted for 17% (n=12,329) of attendances. Patients with dementia comprised 18.9% (n=13,545) of attendances overall, and 22% (n=12,534) of attendances to people aged ≥75.

### Maps

More than 50 chloropleth maps were produced to display the contextual data within the SCAS region of operation, alongside service demand and service outcomes. A selection of the maps considered to be the most relevant to initial discussions was made for the stakeholder sessions.

Initial contextual maps to illustrate the distribution of the population aged ≥65 illustrated the variation in demographics for older adults across the region, with clusters of MSOAs with higher numbers of older adults in the South-West of the region as well as east Hampshire and south Oxfordshire (Figure 1). The most deprived areas are mostly urban settings, including Southampton, Portsmouth, Reading, Slough and Milton Keynes, but there were also rural areas with higher deprivation particularly in South-West Hampshire (the New Forest).

**Figure 1.**
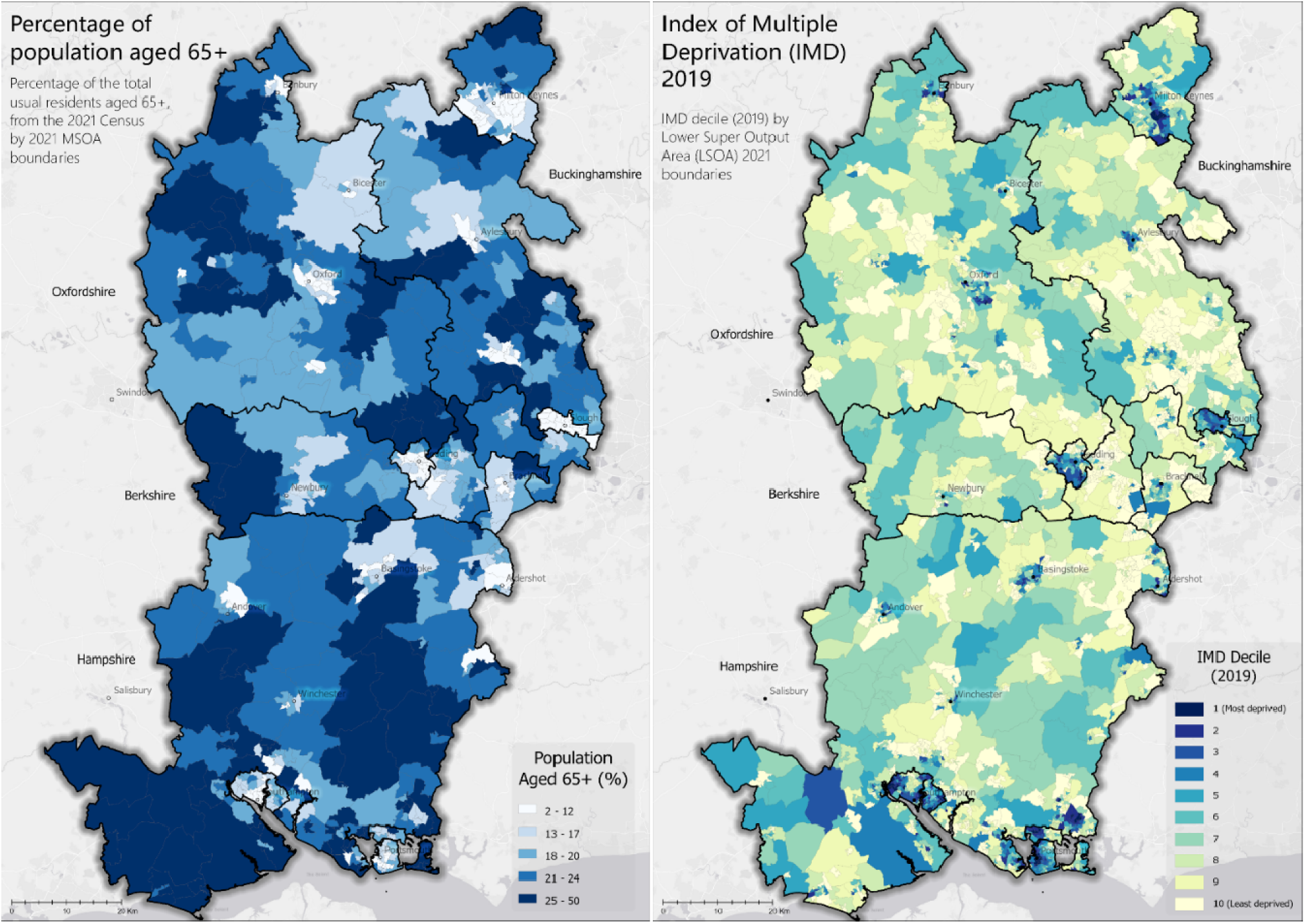
Distribution of older adults and indices of multiple deprivation quintiles in the region.

The maps were an effective way of showing variation in demand in incidents (calls) and attendances across the region (Figure 2). A visual comparison of maps suggests that the distribution of demand mirrored the demographic patterns, and thus highlight areas with higher requirements for ambulance services for patients with potentially complex needs. Patterns of attendance generally reflect the number of incidents, although attendance patterns in some areas were not in line (higher or lower) than would be expected from incident numbers, for example north Oxfordshire and north Hampshire.

**Figure 2.**
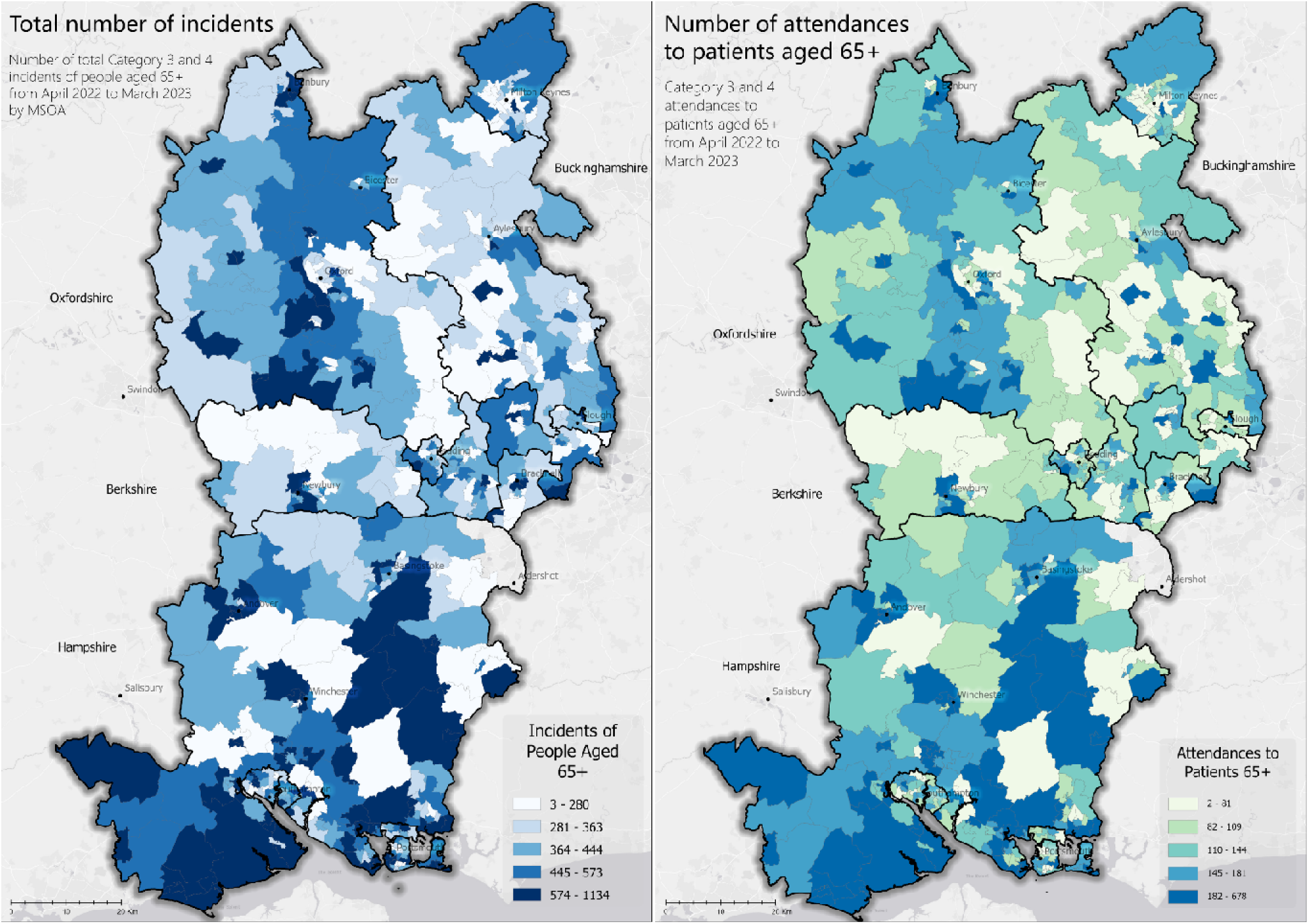
Distribution of 111/999 incidents and ambulance service attendances in the region.

The numbers and proportions of patients conveyed to hospital for all adults aged ≥65 and for patients with a record of dementia in the ePR are shown in Figure 3. Conveyances would usually be taken to an Emergency Department of a local acute care hospital, and this map therefore reflects demand for onward care services. When the numbers of patients conveyed following an attendance are presented as rates, it is evident that there were distinctly different patterns across the region, varying from between 28 to 100% by MSOA. This demonstrates the importance of providing a variety of analyses and outputs, for optimal utility. Areas with particularly high conveyance rates include parts of Buckinghamshire, whilst areas with a higher number of attendances, for example southern Hampshire and north-west Oxfordshire, had lower conveyance rates.

**Figure 3.**
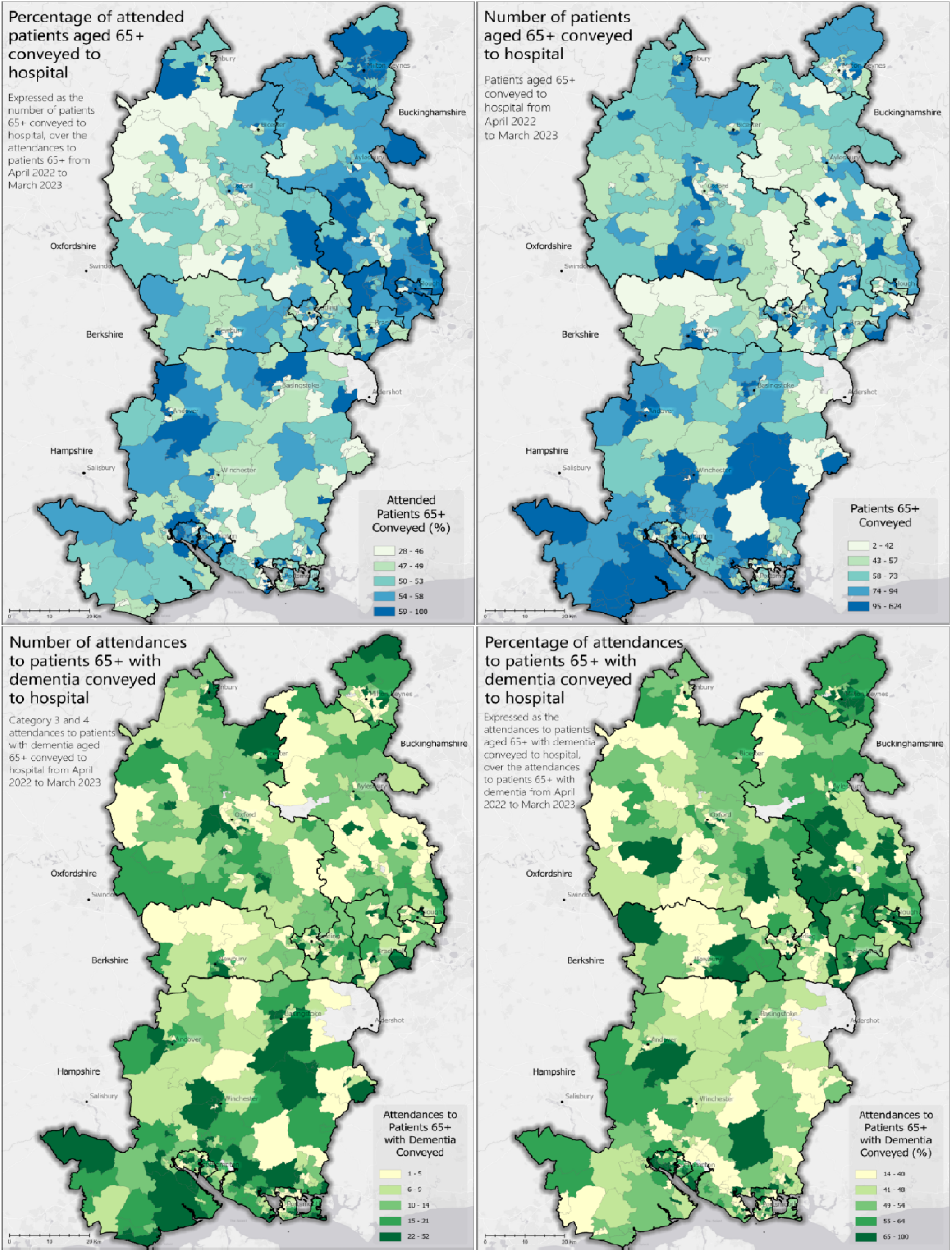
Numbers and proportions of attendances to patients conveyed to hospital.

### Data interpretation and stakeholder views

The discussions in the stakeholder meetings covered four main areas.

1. The usefulness of the maps to identify topics of interest. The group was enthusiastic about the potential of the maps to provide a basis for initial visual scanning of data, which had not previously been looked at for this patient group. The geographical variations in demand and variations in indicators of care provision such as response times demand reflected the group’s experience of delivering emergency care on the ground. The large geographical variations in the rates of conveyances to hospital prompted much discussion, and the maps were considered a good springboard for further conversations to explore operational functions in particular areas.
2. Specific feedback on how to develop the maps. This included incorporating specific locations of SCAS ambulance stations and other key healthcare services such as hospitals, particularly those with services for older people such as frailty assessment centres. Creation of multi-layer maps which incorporated several factors into an interactive map (e.g. population, deprivation and conveyance rates) would be a next step to understand some of the interlinking factors). This information, together with identification of areas with contrasting demand and outcome indicators, was thought to be a good basis for understanding variation in more detail. In this way, a localised whole system can be obtained and in turn may influence localised organisation and delivery of care. Variation due to operational differences between Integrated Care Systems was highlighted as was the resulting impact on SCAS’ service delivery. The exercise provided an opportunity to share good practice throughout the region and to consider strategies to improve equity of service provision.
3. Forming a dissemination plan. Participants identified additional staff members and teams within SCAS with whom it would be useful to discuss the findings and define strategies for future work. This included frontline operational teams in those areas which were outliers from the general patterns shown in the data, and specific teams such as urgent care/falls response teams with particular responsibility for managing older people with complex needs. External stakeholders in the region were also identified in addition to national collaborators with a specific interest in the area for example dementia leads across the ambulance trusts, and quality improvement specialists with a special interest in ambulance services.
4. Capturing ideas for further research or quality improvement. The potential of using geospatial data as a key part of a wider research programme focussing on older people’s emergency care was recognised. Topics of particular interest included: time delays between call/attendance and hospital admissions, particularly for falls; mapping the variation in management between localities against available alternative services; the potential for additional involvement of Community First Responders (CFRs) in the care of complex patients targeted to localities with higher need and higher conveyance rates, and the relevant training and support that they might require; and a focus on the decision-making process around hospital conveyance in different localities, both from the perspective of clinical staff and patients/carers.

A summary of the main points from the two sessions was collated and circulated amongst participants (Supplementary Material, Figure 1).

## Discussion

### Main findings

It is both feasible and acceptable to pursue a collaborative approach between healthcare and academic organisations to analyse data relating to geographical distribution of service demand and outcomes for older people. A multidisciplinary team which includes expertise from business intelligence, geospatial experts, epidemiology, clinical and operational leads is essential to deliver results of relevance to all parties involved. The model of resource and knowledge-sharing between the organisations enabled rapid and efficient project delivery. This was also facilitated by the use of aggregated, non-patient level data in terms of governance and faster approvals.

The maps illustrated significant variations in demand for emergency care services for older adults with non-life-threatening calls across the region, as well as localised variation in subsequent care pathways. These variations are, in turn, likely to impact on outcomes important to patients and follow-on services such as hospitalisation and new admissions into residential care. This initial work has therefore highlighted the potential for use of maps to inform service developments for this patient group both within the ambulance trust and for other health and care partners in the region.

### Lessons learned

This feasibility study facilitated the opportunity for iterative discussions regarding data provision and analysis within the project and for future work. As only a sub-section of the population was included, maps at MSOA level were easier to view and interpret than at LSOA, particularly when focussing on dementia records alone, and the potentially sensitive nature of the data also supported larger group sizes. Understanding the variety of patient pathways, for example between-hospital transfers which were represented in the data was also essential to define a clear baseline population. Prioritisation of resources within already busy health systems is challenging, and supporting work with committed time from academic colleagues with clearly specified joint objectives and benefits is essential to garner management support. Gaining clinical representation from across the operational region is also key to enable data interpretation specific to localities that may be of further interest to explore for this population and to plan next steps. Mapping of underlying care structures is important as healthcare funding still remains inequitable in areas with higher deprivation [20]. Therefore mapping existing health services and stratification of data by deprivation in future work are important, as both of these factors are influenced by locality, and are likely to be key factors influencing conveyance decisions.

### Implications for practice

Although this project was designed to test the feasibility of the workflows and methods, there are already potential implications for practice that can be identified. Having an electronic patient record that effectively captures health conditions common in older people and social factors which may influence their care is key. This enables clinicians on scene to accurately and consistently record information on frailty, dementia and social support. Examining this data on a local area level in conjunction with aggregated datasets from other services, such as local authorities or public health observatories, may provide a more detailed picture of patterns in population need to enable development of solutions for localities with high demand and inequalities in care access and outcomes. Older adults are a key focus for development of alternative care pathways, in accordance with national strategic plans and the SCAS organisational aim of maturing from an emergency point of contact to becoming an integrated and urgent care service. This fosters closer working with local health and care services to continue to prevent inappropriate attendance at emergency departments [21, 22].

The methods of extraction and analyses for this specific dataset can be reproduced to aid evaluation of service developments and shared with the ICS in the region and other ambulance services through the National Ambulance Research Steering Group and the Q Community Ambulance Special Interest Group (SIG), e.g. as a policy brief, as used in previous collaborations [5] to support evidence-based practice and service delivery.

### Implications for further work

The remit and roles of the ambulance service workforce continue to evolve [23], and their intersection with a variety of patient care pathways may be usefully informed by a more precise view of local services, to which geospatial data may contribute. However, feedback from onward care services is rarely received by ambulance clinicians, and with increase in alternative pathways [24], knowledge of patient outcomes may be important to increase confidence in their use [25]. For those aspects of the workforce with focussed geography, such as community first responders, geospatial analysis could be used to inform recruitment initiatives and training programmes to better meet local population needs. Exploring the role of geography in potential differences in transfers to ED from care homes according to Advance Care Plan availability and content may also reveal scope for further locally based interventions [26, 27]. Further development of the ePR to include other data important to inform practice, for example routine assessment of the Rockwood frailty score in older adults and more detailed information on the existing care available at home, would be valuable. Finally, given the evolving changes in population healthcare needs and methods of meeting the demand during and after the global coronavirus health emergency, with concurrent upticks in workforce shortages across health and social care, it is likely that temporal trends in emergency service needs and subsequent conveyances may be observed and therefore geospatial analyses should be regularly reviewed to monitor changes and support timely responses within the integrated care systems.

### Limitations

Although data on acute care locations and services was not added to the maps at this stage, the discussion sessions were useful in identifying this and other data which may influence outcomes and should be considered going forward. Static maps enabling visual comparisons only were used for this feasibility project, but further work can include online maps of increased complexity to overlay variables enabling closer examination of more localised areas within the region, and the ability for the reader to choose which variables are viewed.

### Conclusions

Creation of chloropleth maps to represent older people’s use of ambulance services and their characteristics and outcomes are feasible and desirable. Clarity around data availability and use, governance procedures and involvement of a multidisciplinary team is essential. Maps can highlight areas of variation in care and outcomes and provide a useful starting point to explore equity in service provision both within and beyond urgent care services. Geospatial information could be useful to inform design of prospective studies of care models and also be used in their evaluation.

## Data Availability

The data is the property of the NHS and is not publically available.

## Acknowledgements

We would like to thank the following SCAS staff for their input to the project, discussion sessions and shaping of future work: David Hamer, Jack Ansell, Emily Seear, Andrew Freeman-May, Cheryl Smith, Chris Jackson and Wendy Stonehouse. We are grateful to Martina Brown and Simon Mortimore for providing support and governance oversight from within SCAS.

---

**Supplementary Material – Figure 1.**
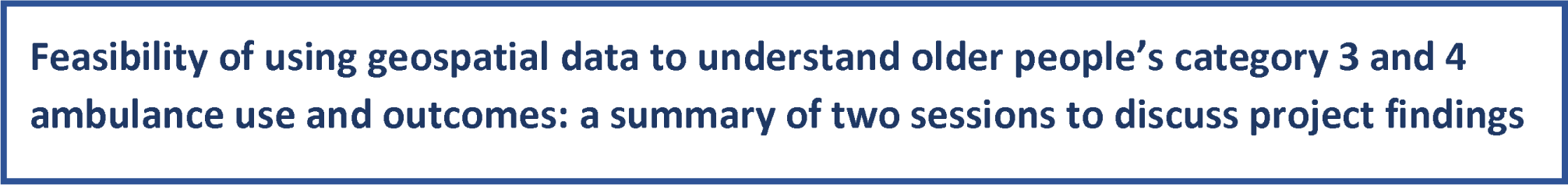
Discussion summary.

## Key messages from slides

➢ There are more older people in certain geographical areas. Areas with high numbers of people aged 65+ include the New Forest, Milton Keynes, Southampton, Aylesbury Vale, Wycombe, Portsmouth.
➢ Patterns of deprivation may be important to consider, as income may influence older people’s access to supportive care at home and healthcare seeking behaviour. Areas of higher income deprivation for older people (IDAOPI) include Slough, Southampton, Reading, Portsmouth, Rushmoor and Milton Keynes.
➢ Areas with higher numbers of category 3 or 4 incidents for people aged 65+ include West Hampshire, Winchester, Andover, the Meon Valley, north Oxfordshire.
➢ Geographical variability in category 3 and 4 mean response times is something to explore further with teams in the area (see maps for distribution of response times).
➢ There is variation in the proportion of attendances conveyed to hospital, with the highest conveyance rates in Slough, Milton Keynes, South Bucks (58% to 61%) and the lowest in West Oxfordshire, Vale of the White Horse and Gosport (45% to 48%).
➢ Numbers of attendances to people with dementia and conveyance rates follow a similar geographical pattern to the overall 65+ population.
➢ There is variability in the distribution of attendances due to falls in people with dementia, with higher proportions in North Hants, south Oxfordshire, mid-Bucks. Some areas have high falls numbers and high conveyance rates – e.g. north New Forest, Newbury South, Milton Keynes.

## How might the maps be useful?

➢ Choropleth maps give a good visual representation of differences. Both ‘numbers’ and ‘proportions’ are useful depending on context.
➢ Maps are good for initial scanning and provide a springboard for discussion. We can then drill down into specific areas for further exploration and resource planning.
➢ The maps can be supplemented by traditional tables of numbers/percentages and more sophisticated statistical analysis.
➢ Maps can be used to identify and share good practice across the region within SCAS and the Integrated Care Systems (ICS) and inform how services available in different areas influence patient pathways.
➢ Can identify areas which have different outcomes (response times, conveyance rates, re-calls within 48 hours) and explore which different strategies are being used in different areas. Can compare similar areas – e.g. urban areas which are near a hospital location, rural areas, areas of similar levels of deprivation or numbers of older people – and see what works.
➢ Can identify areas which could be targeted for modifying or expanding services for the greatest impact – e.g. those with high call numbers (e.g. top 40%) and higher conveyance rates and/or longer response times.
➢ To define which are the best maps to identify differences in demand and potential inequalities in care

## How could the maps be improved?

➢ Include information on locations of other services e.g.:

- falls response vehicles
- locations of hospitals with an Accident and Emergency (A&E) department
- urgent care team locations / Urgent treatment centres (UTCs)
- ambulance stations and stand-by points
- care homes (including breakdown by residential only vs nursing home, and dementia-specialist homes)
- average distance to nearest hospital (miles vs journey time)
➢ Include data (location, availability, use) on referrals to alternative care pathways (e.g. falls service, out-of-hours service, minor injuries unit) and explore correlation with conveyances.
➢ Use 90^th^ centile for response times in addition to the average to get an idea of range.
➢ May also need category 1 & 2 call numbers to understand overall picture and where the category 3 & 4 calls may be more likely to have delays – i.e. overall service flow.

## Which other stakeholders should we discuss this with?

➢ Further discussion within SCAS to contextualise data and prioritise further work - involve operational leads, Community First Responders (CFRs), road staff from different locations across the region (e.g. contrasting areas such as New Forest, Milton Keynes), urgent care/falls response teams, clinical service desk that supports the CFRs, pathways team.
➢ Disseminate findings and proposed future directions within SCAS: Board meetings, Clinical Review Group.
➢ Wider regional stakeholders: ICSs, community health Trusts, GPs, community nurses, palliative care/hospice at home teams, adult social care, care home staff.
➢ National collaborators: Dementia leads across ambulance trusts, Health Foundation Q Community Ambulance Special Interest Group, Welsh Ambulance Service.
➢ University networks and opportunities e.g. geospatial network, ARC Wessex.

## Potential areas for further Research/Quality Improvement

➢ Hospital admissions due to “long lie” following a fall – how many are avoidable due to time on floor waiting for response? How does this vary between areas? What can different areas learn from each other – e.g. in relation to other services available, configuration/availability of urgent care teams, CFR training/capacity, non-hospital referrals.
➢ Is there a greater role for CFRs in category 3 or 4 calls for older people that may reduce response times and number of conveyances to hospital?? E.g. using the ‘eyes on scene’
➢ Exploring use of alternative referral pathways, e.g. differences in availability and use according to geography, and how this impacts on response times and conveyance.
➢ Explore experiences of CFRs in attending non-injury falls.
➢ Staff experience of attending patients with dementia – decision-making and relation to levels of clinical experience/confidence/training.
➢ Patient/carer experience of being attended by a CFR first vs waiting for paramedic team – benefits and drawbacks from their perspective.
➢ Explore patient/carer decision making around being taken to hospital - variation according to patient characteristics, how this ties in with the clinical staff judgement, consequences (e.g. re-calls within 48 hours).

